# The impact of social protection interventions on treatment and socioeconomic outcomes of tuberculosis-affected people and households in low income, high burden settings: A systematic review and meta-analysis

**DOI:** 10.1101/2025.03.04.25323276

**Authors:** Mollie Hudson, Heather Todd, Talemwa Nalugwa, Ann Schraufnagel, Canice Christian, Delia Boccia, Tom Wingfield, Priya B. Shete

**Author notes:** Corresponding author: Mollie Hudson.

## Abstract

**Introduction:** Tuberculosis (TB) is the leading cause of death due to infectious disease worldwide. Social protection interventions can benefit TB-affected households. We conducted a systematic review and meta-analysis to quantify the effectiveness of social protection on TB treatment and socioeconomic outcomes.

**Methods:** We identified articles published from January 2012 to July 2024 by searching PubMed (includes MEDLINE), Embase, and Web of Science. We included studies that described at least one social protection intervention and reported on either TB treatment or socioeconomic outcomes for people with TB or TB-affected households. Random-effects meta-analysis was used for our primary outcome of interest, TB treatment success (treatment completion or cure). We performed a meta-regression to evaluate the association of study characteristics with odds of TB treatment success. Risk of bias was assessed using the Newcastle Ottawa Scale and the Cochrane Risk of Bias tool. This review was registered prospectively in the PROSPERO database (registration number CRD42022382181).

**Findings:** Our search generated 47,245 articles. Of the 50 which were eligible for inclusion, 36 reported TB treatment outcomes, 8 reported on socioeconomic, and two studies reported both TB treatment and socioeconomic outcomes. Random-effects meta-analysis of 24 articles found that people with TB who received social protection interventions during treatment had 2.23 times the odds of TB treatment success (95% CI 1.82, 2.74, I^2^ 93.8%).

**Conclusion:** Social protection interventions significantly improve odds of TB treatment success. Outcomes and definitions used in our study have the potential to guide further research and implementation of social protection for TB-affected populations.

**Summary Box:** *What is already known on this topic:* Several studies have found that social and financial interventions designed to mitigate socioeconomic risk and promote resiliency, termed social protection interventions, have the potential to improve treatment outcomes for tuberculosis (TB), including treatment completion and cure. Additionally, several studies have demonstrated that social protection interventions can improve socioeconomic outcomes among TB-affected households such as averting catastrophic costs and negative financial coping strategies.

*What this study adds:* This is the first systematic review and meta-analysis that comprehensively evaluates the impact of TB specific and TB sensitive social protection interventions on both TB treatment and socioeconomic outcomes, thereby generating evidence on the ability of these interventions to curb the well-known cycle of TB disease and poverty. Through the use of an extensive list of search terms, expanded and systematic inclusion of outcomes of interest, and a focused definition of social protection interventions, our systematic review included the adequate number of high-quality studies needed to conduct a meta-analysis. Additionally, our systematic review evaluated implementation outcomes described in eligible studies which provides the basis for feasibility of these strategies in programmatic settings.

*How this study might affect research, practice or policy:* Our study provides evidence that social protection interventions, when used in conjunction with standard biomedical treatment, have the potential to significantly improve TB treatment outcomes. This study fills an essential gap in existing synthesized evidence of the impact of social protection interventions on TB, socioeconomic, and implementation outcomes. Our findings also highlight the need for standardized definitions of social protection, as well as uniform reporting procedures, to better help evaluate the impact of social protection interventions for TB-affected individuals and households. Addressing these gaps provides scientific basis for meeting the commitments articulated in the 2023 United Nations General Assembly high level meeting for TB which calls for social protection for all individuals with TB.

## Introduction

Tuberculosis (TB) is one of the leading causes of infectious disease deaths worldwide. Despite effective and widely available treatment, an estimated 10.8 million people were infected with TB and 1.25 million people died from TB in 2023.^1^ TB-affected individuals are often trapped in a vicious cycle of poverty; impoverished individuals often have risk factors that make them more susceptible to TB (e.g. crowded living conditions, poor access to care, malnutrition), and becoming ill with TB often precipitates devastating economic consequences incurred from the such as costs associated with of care-seeking and treatment and lost wages due to missed work.^2,3^ TB-affected households often use negative financial coping strategies and experience catastrophic costs, defined as total costs in excess of 20% of the annual household income.^4^

To break out of the cycle of poverty and disease, interventions are urgently needed that address both underlying social and economic determinants and vulnerabilities of TB-affected individuals. Social protection interventions, broadly defined by the World Bank as systems that “help the poor and vulnerable cope with crisis and shocks, invest in the health and education of their children, and protect the aging population,”^5^ are such a strategy. Social protection interventions are a key pillar of the World Health Organization (WHO) End TB Strategy, the United Nations’ Sustainable Development Goals (SDGs), and the UN Declaration on TB.^6,7,8^ These interventions include cash transfers to eligible populations, job training programs to support income generation, and nutritional or food security programs. When implemented effectively, social protection can decrease TB incidence, improve TB treatment outcomes, and improve socioeconomic outcomes.^3,9,10^

While prior studies, including some systematic reviews, have found that social protection interventions can improve TB treatment outcomes,^11^ they were limited in their ability to widely inform policy and programmatic implementation of social protections due to more narrow focus on specific types social protection interventions, or alternatively, including interventions not defined as social protections by normative institutions such as the World Bank. No systematic review has described the impact of social protection interventions on socioeconomic outcomes, including catastrophic costs, dissaving, or standardized measures of poverty. The objective of this systematic review and meta-analysis is to provide synthesized evidence of health and socioeconomic effectiveness of social protection interventions for TB-affected individuals and households required to inform global scale-up of these strategies.

## Methods

We used the Population, Intervention, Comparison, Outcome, and Time (PICOT) format to define our research questions for this systematic review and meta-analysis (**Appendix A**):

1. Do people with TB who have enrolled in at least one social protection intervention demonstrate an improvement in TB treatment success (treatment completion or cure) compared to people with TB who have not enrolled in and/or been recipients of social protection interventions?
2. Do people with TB who have enrolled in at least one social protection intervention have better socioeconomic outcomes, including lower rates of catastrophic costs and dissaving, compared to people with TB who have not enrolled in and/or been recipients of social protection interventions?

This systematic review protocol was guided by the Preferred Reporting Items for Systematic Reviews and Meta-analysis (PRISMA) protocol checklist.^12^ A scoping review^13^ was also conducted to landscape the available evidence and to inform the protocol for this analysis, which has been published previously.^14^

### Search strategy and selection criteria

Our initial search was conducted March 2021, and then repeated September 2023 and January 2025 to include articles published in English through July 2024. We included three electronic databases: PubMed (includes MEDLINE), Embase, and Web of Science. We used Google Scholar Advanced to search selected, relevant databases, explained in detail in our scoping review.^13^ Articles were imported into Covidence^15^ for systematic screening by the study team (**Appendix B**) in accordance with PRISMA guidelines. Articles deemed eligible for inclusion were reviewed by three researchers (MH, HT and CC; **Appendix B**).

We included randomized controlled trials, cross-sectional and cohort studies. Studies were included if they described people with pulmonary and extra pulmonary TB, people with drug-sensitive (DS-TB) and drug-resistant (DR-TB)/multi-drug resistant TB (MDR-TB), people either with or without HIV-TB co-infection, or TB-affected households in either low-to-middle-income countries (LMICs) and/or high burden TB countries as defined by WHO or the World Bank.^1,16^ We only included studies published since 2012, which coincided with the “World Bank’s Social Protection and Labour Strategy 2012-2022,”^17^ which supported initiatives on reducing socioeconomic risk, after which definitions of social protection were expected to be uniform.

### Intervention

We only included studies in which the main independent variable was enrollment in, and/or receipt of, a social protection intervention as defined by the World Bank.^5^ We included social protection interventions specifically designed for individuals or households affected by TB (*TB-specific social protection interventions*), for which microbiologic or clinical diagnosis of TB is a requirement of eligibility for the social protection intervention, as well as social protection interventions designed for individuals or households inclusive of those affected by TB but identified as eligible based on non-TB criteria (*TB-sensitive social protection interventions*).^18^

### Outcomes

We only included studies for which the main dependent variable was at least one standardized outcome related to TB treatment including cure, treatment completion, treatment success (a composite variable of TB cure and treatment completion) mortality/death, treatment default, loss to follow up,^1^ and/or one standardized socioeconomic outcomes such as catastrophic costs, dissaving or impoverishment up, in accordance with global definitions.^1^

### Screening and descriptive analysis

Titles and abstract were systematically and independently screened by the study team (Appendix B). Potentially eligible articles underwent full text review. Questions about study inclusion were discussed by reviewers (MH, HT) with adjudication by third reviewer (TN) or senior researchers (TW, PBS). **Figure 1** describes the search process in accordance with PRISMA protocols for systematic reviews.

**Figure 1.**
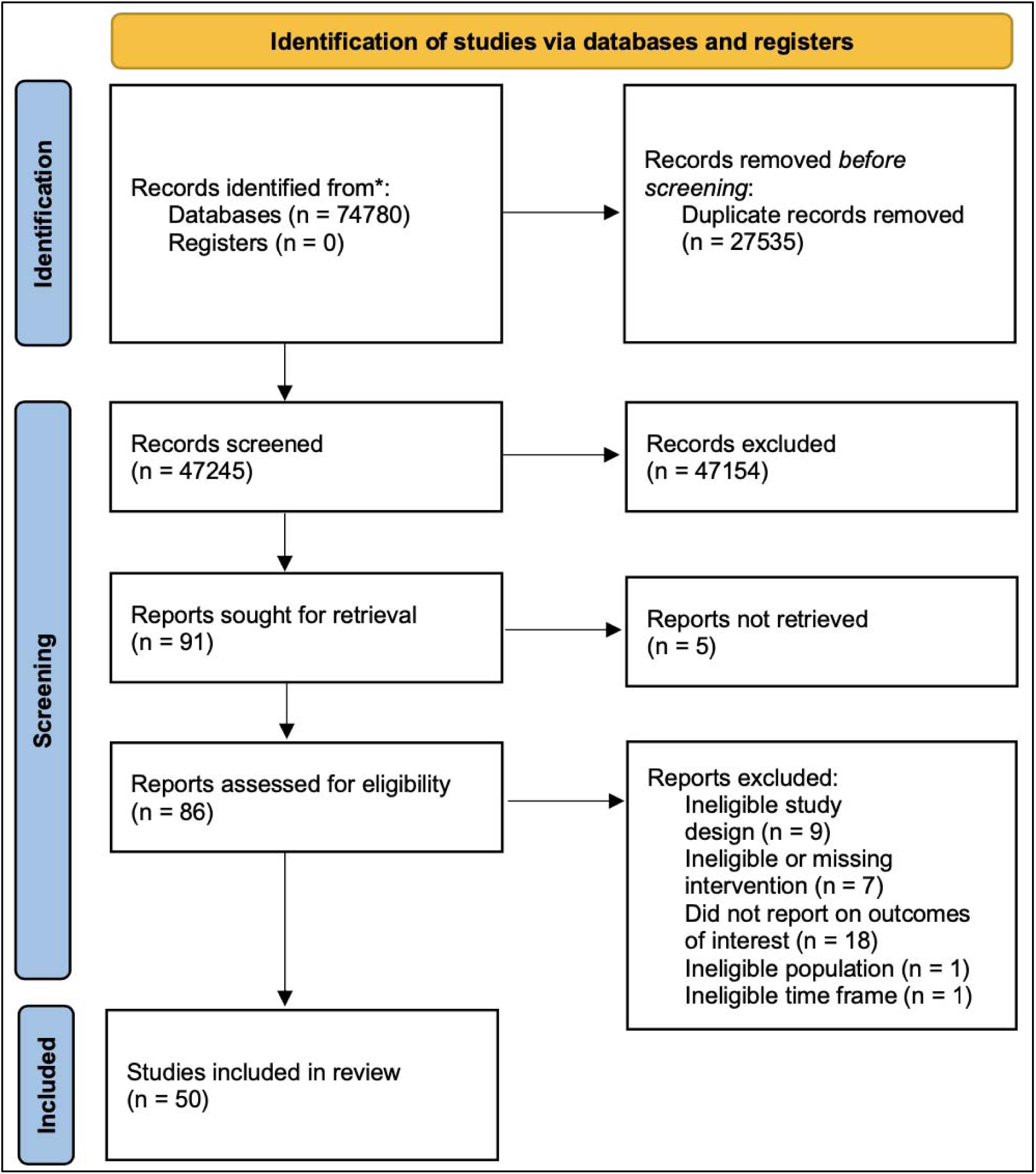
Flow diagram of search process to identify eligible studies using PRISMA.

### Quantitative analysis

Three reviewers (MH, AS, CC) independently extracted quantitative data reported in eligible papers (**Supplementary Tables 1-4**). For the meta-analysis, authors extracted data describing the number or percent of individuals who attained TB treatment success in both the treatment (receipt of social protection in conjunction with standard biomedical treatment) and the control (standard biomedical treatment only) groups. All estimates of effect for dichotomous outcomes (e.g. “achieved treatment success” versus “did not achieve treatment success”) were reported as odds ratios with a 95% confidence interval. We used a random effects model^19^ for our meta-analysis to account for heterogeneity between studies. All calculations were conducted in STATA BE version 17.^20^

A logistic meta-regression was performed to evaluate the association of study characteristics with the odds of TB treatment success. We selected a limited number of study characteristics to not overfit the model including: 1) study quality, 2) whether the study included individuals with MDR-TB, and 3) study setting. Study quality for studies scored using the Newcastle Ottawa Scale (NOS); a summed score of 9-8 was categorized a high-quality study; a summed score of 7-4 (inclusive) was categorized as a medium quality study, and a summed score of 3-0 (inclusive) was categorized as a low-quality study. Risk of bias assessments are summarized in **Supplementary Tables 5 and 6**.

### Qualitative analysis

We reviewed qualitative manuscripts according to theme and content, and only included papers that described either TB treatment or socioeconomic outcomes. Findings are summarized in **Supplementary Table 7**.

### Risk of bias in individual studies

We used the Cochrane Risk of Bias (RoB) tool^21^ for RCTs and the Newcastle Ottawa Scale^22^ (NOS) for all studies that quantitatively reported on outcomes. Studies are scored in 3 areas – selection (scored 0-4) comparability (scored 0-2), and outcome (scored 0-3), with higher scores indicating a positive assessment (less likely to be biased). Risk of bias was appraised by three researchers independently (MH, HT, AS).

## Results

After removing duplicates, we screened 47,245 titles and abstracts (**Figure 1**). After removing 44,316 articles based on title and abstracts, we evaluated 86 articles for eligibility. Thirty-six articles did not meet our criteria for eligibility (**Supplementary Table 7**). Of the 50 included articles, 46 articles reported primarily on quantitative outcomes, while four articles reported only on qualitative outcomes.^23–26^ The results of our search are reported in **Figure 1** in accordance with PRISMA guidelines. Study characteristics including interventions, outcomes, and setting are summarized in **Supplementary Tables 1-4**.

Of the 46 articles that reported on quantitative outcomes, 38 (83%) were cohort studies, two (4.8%) were randomized controlled trials, three studies (6.5%) were cross sectional, one study and three studies (6.5%) were cluster randomized trials. No studies were quasi-experimental or ecological. Studies were conducted across 18 different countries (**Supplementary Tables 1 & 2)** and across urban, rural, and mixed settings (**Supplementary Tables 1 & 2)**. Six different categories of social protection interventions were described in the studies that reported quantitative outcomes **(Supplementary Table 3**), with cash transfers (n=22, 48%) most frequently described. Social protection interventions described in the qualitative studies reviewed (n=4) included psychosocial, nutritional, and financial support in the form of transportation stipends. Interventions were described as either TB sensitive, e.g. eligibility criteria primarily pertaining to measures of poverty, or TB specific, for which only TB affected individuals and/or households were eligible. Thirty studies reported TB treatment success as the primary outcome of interest. Other outcomes included were mortality, TB treatment default (treatment interruption of at least two months^1^), loss to follow up, treatment failure, or a combination of TB treatment outcomes (**Supplementary Table 4**).^27^

Nine studies reported primarily on socioeconomic outcomes, while two studies reported secondary socioeconomic outcomes (**Supplementary Table 4**). Five papers described how social protection reduced the risk of incurring catastrophic costs.^3,28–31^ As a secondary outcome, one study found that expenses related to hospital admissions decreased as a result of social protection.^32^ Three studies found that social protection did not improve outcomes,^33–35^ while one study found that outpatient TB treatment outcomes improved with social protection while inpatient TB treatment outcomes were unchanged.^36^ Most studies did not describe implementation process metrics, namely fidelity, reach, and coverage (**Supplementary Table 4**). Reasons for poor implementation fidelity reported by nine studies were related to administrative issues such as inadequate banking systems and/or delayed or non-disbursement of social protection benefits.

Random-effects meta-analysis was used for our primary outcome of interest, TB treatment success. Of the 30 studies that quantitatively reported on TB treatment success, two studies did not have a control group^36,37^ and two studies did not report findings such that data could be extracted for meta-analysis calculations.^38,39^ One study did not provide information about how the historical cohort was recruited and compared to the intervention cohort.^40^ Lastly, although Wrohan et al.^41^ reported treatment success by social health insurance status, researchers only evaluated one type of social health insurance as the “social protection intervention,” and did not include additional information about the cohort of individuals not using social health insurance. Authors of this meta-analysis agreed that there was not sufficient information provided in the study to distinguish the intervention from the control cohort. We therefore included 24 studies in our meta-analysis. Lastly, one study included in the meta-analysis described how although a larger cohort of individuals was enrolled in a social protection intervention, only 233 individuals actually received the disbursed funds.^42^ Authors therefore modified their analytic approach according to how many individuals actually received disbursed funds; ^42^thus, we included only those who were enrolled and received the intervention in our meta-analysis.

People with TB who were exposed to social protection interventions in conjunction with standard biomedical treatment had approximately two times the odds of achieving TB treatment success (**Figure 2**). The distribution of study effects is relatively narrow, with almost all studies reporting a positive effect. Our pooled estimate has an odds ratio of 2.23 with a confidence interval of 1.82 to 2.74, suggesting moderate heterogeneity, even with an I^2^ of 93.75%.

**Figure 2.**
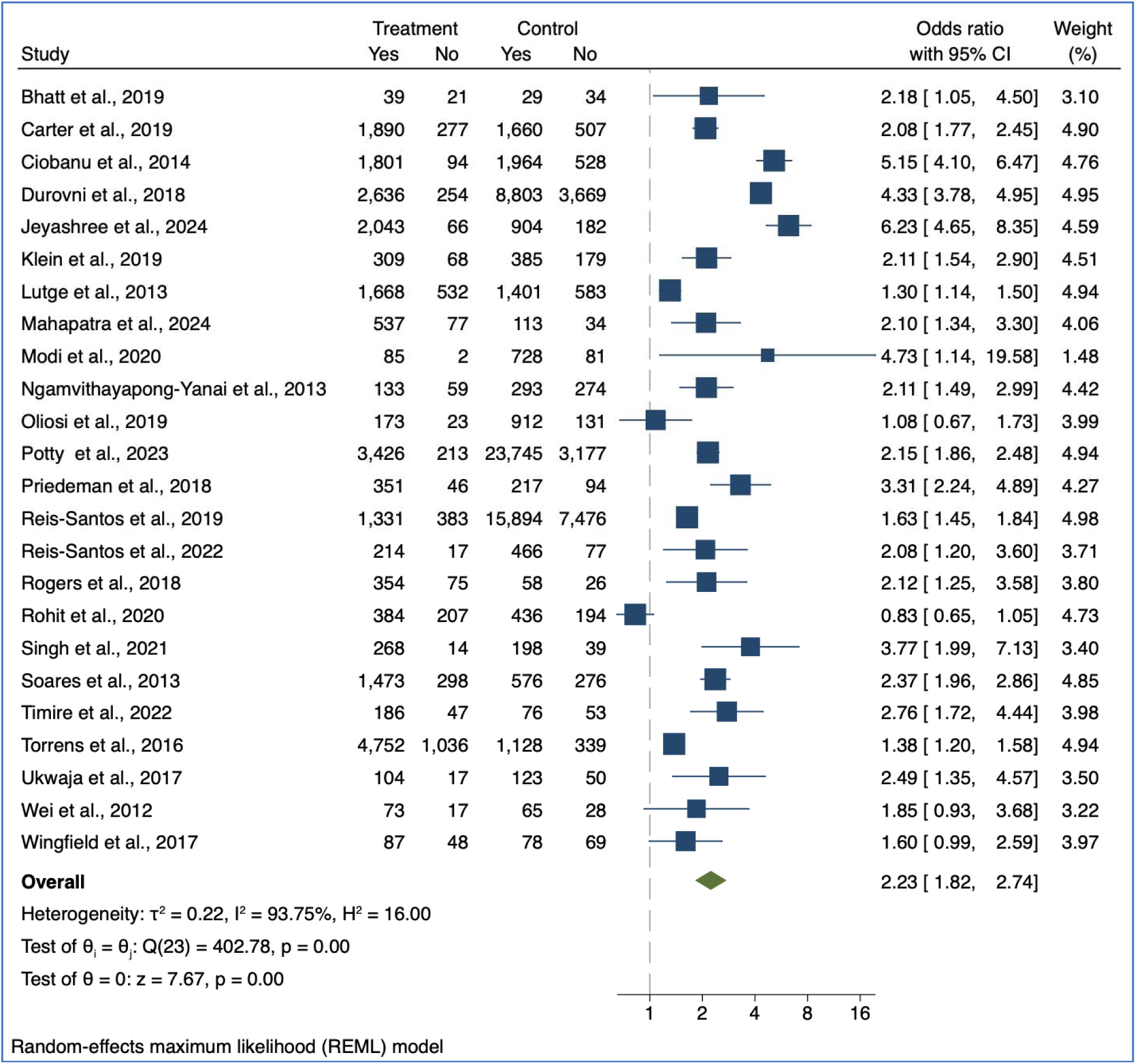
Meta-analysis of treatment success rates in individuals who were enrolled and/or recipients of social protection vs. those who were not enrolled and/or recipients of to a social protection intervention. Treatment=exposed to social protection, control=not exposed to social protection. The two RCTs included in the meta-analysis were Reis-Santos et al., (2019) and Wingfield et al. (2017).

Results of the risk of bias assessments are described in **Supplementary Table 5** and **Supplementary Table 6**. AS, CC and MH used the Cochrane RoB Tool for Randomized studies,^43^ and the NOS^44^ to independently evaluate risk of bias in non-randomized studies that reported quantitatively on outcomes. We found the quality of the studies included to be high based on criteria specified in the Cochrane RoB tool and the NOS.

The odds of TB treatment success were regressed on study covariates as shown in **Table 1**. Conducting a study in a rural setting, the inclusion of MDR-TB, and a non-cash social protection intervention type were all not significantly associated with lower study odds of TB treatment success. Adjusting for other variables, the relationship between study quality and study odds ratio was not statistically significant, there was a trend for lower odds ratios of treatment success in high quality (1.9 [−5.4, 1.7]) and medium quality (−2.3 [−7.5, 3.1]) studies compared to low quality studies.

**Table 1.** Regression coefficients and p values from meta-regression of the study odds of TB treatment success across covariates of interest (n=24)

| Variable | Regression coefficient (95% CI) | P value |
| --- | --- | --- |
| Study quality |  |  |
| *Low quality (reference)* | -- | -- |
| *Medium quality* | -2.3 (-7.5, 3.1) | 0.4 |
| *High quality* | -1.9 (-5.4, 1.7) | 0.3 |
| Inclusion of individuals with MDR TB |  |  |
| Yes | 0.9 (-1.7, 3.4) | 0.5 |
| No | 0.9 (-1.9, 3.7) | 0.5 |
| Study setting |  |  |
| *Primarily rural* | -.5 (-5.5, 4.6) | 0.8 |
| *Mixed urban/rural* | -.3 (-3.1, 2.8) | 0.8 |
| *Primarily urban* | -.6 (-3.9, 2.6) | 0.7 |
| Social protection type |  |  |
| *Cash (reference)* | -- | -- |
| *Nutrition* | .6 (-2.3, 3.5) | 0.7 |
| *Psychosocial* | -.2 (-3.1, 2.6) | 0.9 |
| *Mix of interventions* | .3 (-1.8, 2.4) | 0.8 |

Four eligible studies utilized qualitative methods to describe the impact of social protection interventions. While the number of studies were too few to provide a meaningful content analysis, we identified themes across studies. Primarily, we evaluated qualitative studies according to type of social protection intervention and actual or perceived success or non-benefit of an intervention. We found that social protection interventions were generally had high levels of acceptability among health care workers and people affected by TB, ^23,24,45^ and that interventions that offset the costs of direct non-medical costs related to TB enabled treatment adherence. ^45,46^ However, no studies explicitly evaluated potential negative perceptions or barriers to social protection.

## Discussion

The results of this systematic review and meta-analysis provide a holistic assessment of the evidence available on the effectiveness of social protection interventions on TB treatment outcomes. Where available, we also synthesized socioeconomic and implementation outcomes of these interventions. Our search yielded 50 studies overall, including 24 studies eligible for a meta-analysis demonstrating that social protection inventions, when administered to individuals affected by TB and in conjunction with standard of care treatment, can double the odds of TB treatment success. This finding suggests that social protection interventions can dramatically improve TB treatment outcomes for TB affected individuals. These results support the need to scale-up these programs globally in keeping with recent UN General Assembly High Level meeting commitments to the fight against TB which calls for all TB-affected individuals have access to social protection interventions to prevent financial hardship by 2027.^7^

Our findings demonstrated similar results to prior systematic reviews. For example, a 2018 study conducted by Richterman et al.^11^ found that cash transfer interventions for TB-affected individuals was associated with 1.77 times the odds of a “positive TB treatment outcome.”^11^ A 2018 systematic review and meta-analysis by Andrade et al. found that participants who were beneficiaries of social protection interventions had 1.09 times the odds of TB treatment success and 1.11 times the odds of a cure compared to those who were not beneficiaries of social protection interventions.^47^ While these prior studies are informative, we aimed to be more expansive and consistent in the definitions used for this review to align with global guidance on implementation of social protection for TB-affected households.^48^ In particular, our analysis was based on current comprehensive definitions of social protection as endorsed by the World Bank and International Labor Organization (**Appendix C**).^5,49^ Although this search strategy yielded many articles that did not meet eligibility criteria, we captured studies that we may not have otherwise found with a narrower definition of social protection.

Our study also sought to estimate the potential impact of social protection interventions on reducing the risk of catastrophic and out-of-pocket costs among TB-affected households ^3,23,28^ However, the types of socioeconomic outcomes reported on by eligible studies were variable and not comprehensive. For example, only five studies reported on the socioeconomic outcomes we specified in our search criteria; no studies reported on impoverishment measures. The limitations of these findings highlight the need for standardizing socioeconomic outcomes for future studies in a manner that supports programmatic monitoring and evaluation of global indicators.

Operational challenges to providing social protection feasibly and sustainably for TB-affected individuals have long been cited as a barrier for program coverage and uptake.^50^ We analyzed eligible studies to inform operational and implementation outcomes including feasibility, coverage, or uptake. Unfortunately, the majority of studies did not describe these implementation outcomes or did not use standardized metrics (**Supplementary Table 4**). Of studies which reported implementation challenges, several describe administrative hurdles to provision of benefits; for example, unreliable banking systems resulting in delayed or failed disbursement of funds to beneficiaries and mitigating the potential impact of the intervention (**Supplementary Table 4**). Additional research to identify barriers and facilitators to accessing social protection, as well as standardizing and reporting implementation outcomes, would provide needed evidence on feasibility, acceptability, and coverage necessary to support the deployment, monitoring, and scale up of interventions.

While the methods utilized in this systematic review, including robust risk of bias assessment and meta-regression, enhance our findings, our approach has limitations. The majority of studies that met eligibility criteria were from middle income settings, primarily Brazil, limiting generalizability of our findings; half of the high burden TB countries are located in Africa, are predominantly classified as low-income, and lack the robust landscape of existing social protection programs as Brazil.^1^ Second, the majority of studies reported cash transfer interventions. Additional evidence about other types of social protection interventions would be beneficial, especially as National TB Programs and other key stakeholders begin designing, implementing, and scaling up interventions.

In conclusion, social protection interventions can significantly improve TB treatment outcomes. In our meta-analysis, individuals who had access to social protection interventions had twice the odds of attaining TB treatment success compared to those who did not have access to social protection interventions. Our results, which are grounded in high quality evidence and robust to bias based on meta-regression, also suggest that social protection improved other outcomes, such as decreased mortality and treatment default in addition to potential socioeconomic outcomes. The standardized outcomes and definitions used in this systematic review and meta-analysis have the potential to guide further research on social protection programs for TB-affected populations.

## Data Availability

All data produced in the present work are contained in the manuscript.

## Contributors

Author contributions are described in **Appendix B**. All authors had full access to all of the data in the study and had final responsibility for the decision to submit for publication.

## Data sharing

No original data was collected for this study. All data included was extracted from peer reviewed, published studies.

## Patient and public involvement

It was not appropriate or possible to involve patients or the public in the design, or conduct, or reporting, or dissemination plans of our research.

## Acknowledgements

Funding for this study comes from the Nina Ireland Program in Lung Health (PI: Shete, fund number 7710-138404-7504523-45).

## Appendix A PICOT framework

| **PICOT statement** | **Population** | **Intervention** | **Comparison** | **Outcome** | **Time** |
| --- | --- | --- | --- | --- | --- |
| #1 | TB-affected individuals and households | Social protection in conjunction with standard biomedical TB treatment (medication to treat TB, standard medical appointments) | Individuals who are not recipients of/enrolled in social protection; biomedical TB treatment only | TB treatment outcomes (cure, treatment completion, death, treatment default, TB treatment success) | 2012-2023 |
| #2 | TB-affected individuals and households | Social protection in conjunction with standard biomedical TB treatment (medication to treat TB, standard medical appointments) | Individuals who are not recipients of/enrolled in social protection; biomedical TB treatment only | Improved socioeconomic outcomes (e.g. reduced catastrophic costs, dissaving, various measures of poverty) | 2012-2023 |

## Appendix B Study team

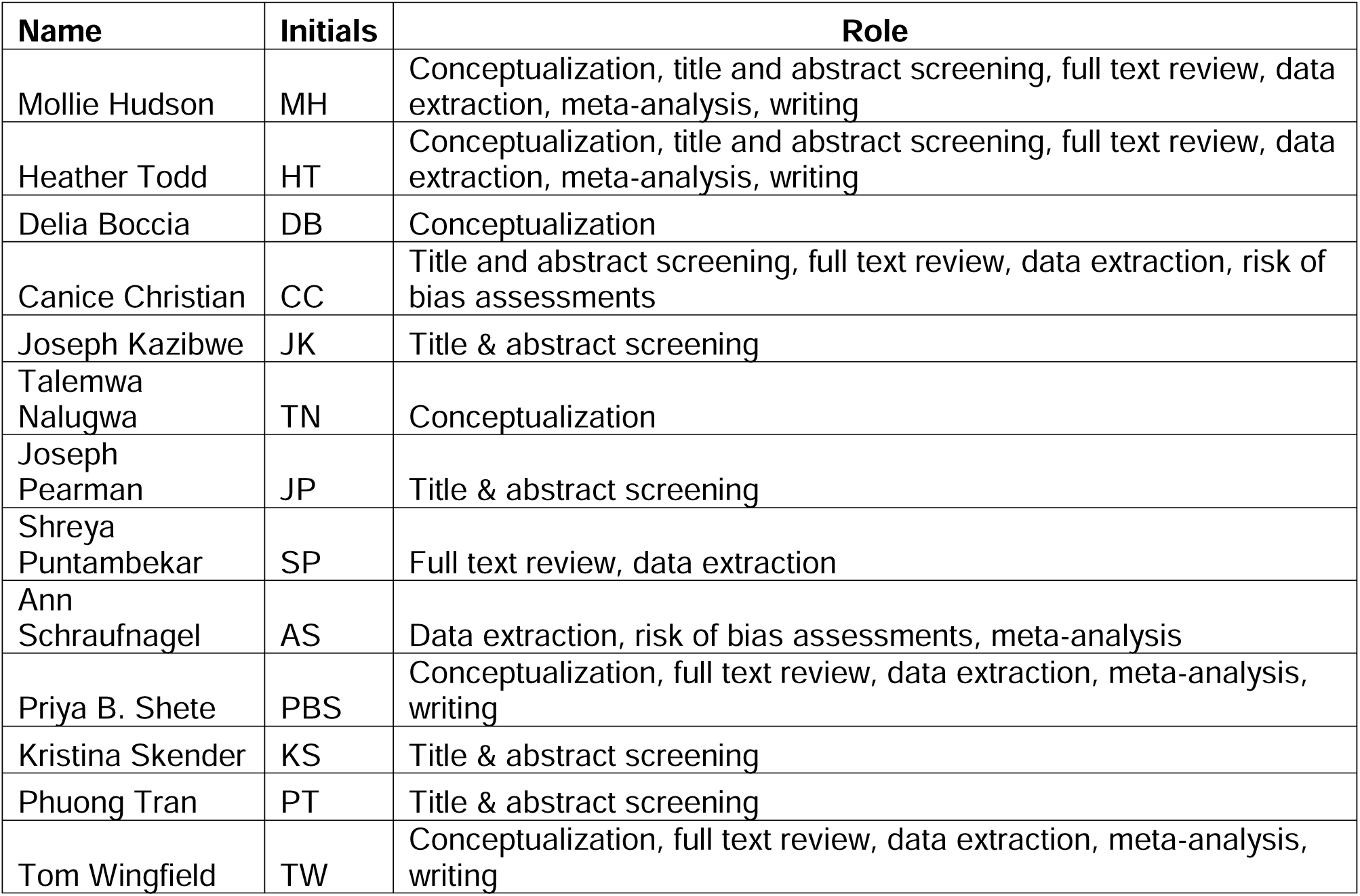

## Appendix C Outcomes by PICOT

- **Outcomes for PICOT #1:** Primary and secondary outcomes related to TB treatment and catastrophic costs, and the nature of the social protection intervention.

- Primary TB treatment outcome:

- TB treatment success
- Death
- Secondary TB treatment outcomes:

- Cure
- Treatment completion
- Adverse TB treatment outcomes:
- Loss to follow up
- Relapse
- Treatment failure
- While this terminology as no longer used, it is likely that studies will have used this terminology.
- No evaluation
- Outcomes for PICOT #2:

- Catastrophic costs

- Catastrophic costs (total costs of entire TB illness >20% of the same household’s annual pre-TB income)
- Costs
- Direct medical
- Direct non-medical
- Indirect (lost income, time, and productivity)
- Of note, these metrics may be calculated different based on the study approach, which will have to be taken into account when analyzing our findings.
- Dissaving

- Dissaving
- If the patient/household took out a formal or informal loan
- If the patient/household sold an asset or item
- If the patient/household used savings
- If the patient/household took a child out of school
- Reduced household food consumption
- Percent poor based on multidimensional poverty index scores (reference: World Bank Group. *Reversals of Fortune: Poverty and Shared Prosperity 2020*.; 2020. doi:10.1038/302765a0)

- Percent poorer than median poverty score (person with TB and/or TB-affected household)
- Experiencing extreme poverty
- Below specified higher poverty lines (USD $3.20 or $5.50 (TB-affected household)
- % below SPL (TB-affected household)
- Person with TB and/or TB-affected household’s perception of poverty and the impact of TB on their poverty
- For example, if a study used the WHO TB Patient Cost Survey, which asks questions about how TB illness has affected individual and/or household level poverty

## Appendix D Search strategy keywords

Appendix D: Search strategy keywords
|  | Generic keywords | Other keywords |
| --- | --- | --- |
| 1 | Tuberculosis ("TB treatment terms") | TB, Mycobacterium tuberculosis, pulmonary TB, TB-affected, TB-infected, TB patients, drug-resistant TB, TB individuals/households, TB prevalent, TB cases Pulmonary TB, PTB |
| 2 | social protection ("intervention terms") | Social safety net<br>Socioeconomic support<br>Social support<br>Economic support<br>Financial support<br>Cash transfers; food-based programs, supplementary feeding programmes, food stamps, vouchers, and coupons; in-kind transfers such as school supplies and uniforms; conditional cash transfers; price subsidies for food, electricity, or public transport; public works programmes; and fee waivers and exemptions for health care, schooling, and utilities, welfare<br>Food baskets, food rations<br>Protections against shocks<br>Social risk management<br>Transportation<br>Government financing<br>Reimbursement<br>Low and middle income, LMIC<br>Support groups, education, community support |
| 3 | Support ("intervention terms") | Intervention, incentive, program, scheme, policy, assistance, livelihood support, enabler |
| 4 | Impact ("outcome terms") | Affect, effect, association, associated, consequence |
| 5 | Treatment ("outcome terms") | Outcome, success, rates, unsuccessful, uptake, enrolment, adherence, cured, completed, treated, follow-up, loss to follow-up, relapse, recurrence, adverse outcome, diagnostic pathways, TB testing, quality of life, default, care cascade |
| 6 | Socioeconomic ("outcome terms") | Outcome, financial burden, economic burden, economic consequences, social consequences, socioeconomic consequences, social impact, socioeconomic impact, costs, expenditure, |
|  |  | expenses, spending, catastrophic expenditure, catastrophic costs, impoverishment, coping strategies, poverty, food security, loans, sold assets, dissaving, deprivation, defray, mitigate |

## Appendix E Search strategies

Appendix E: Search strategies
|  | Searches | Results |
| --- | --- | --- |
|  | (Tuberculosis[Title/Abstract] OR TB[Title/Abstract] OR "Mycobacterium tuberculosis" [Title/Abstract] OR "Pulmonary TB" [Title/Abstract] OR "TB-affected" [Title/Abstract] OR "TB infected" [Title/Abstract] OR "TB patients" [Title/Abstract] OR "Drug-resistant TB" [Title/Abstract] OR "TB individuals" [Title/Abstract] OR "TB-affected households" [Title/Abstract] OR "TB prevalent" [Title/Abstract] OR "Pulmonary tuberculosis" [Title/Abstract] OR "Pulmonary TB" [Title/Abstract] OR PTB[Title/Abstract]) AND (Social protection[Title/Abstract] OR "Social safety net" [Title/Abstract] OR "Socioeconomic support" [Title/Abstract] OR "Social support" [Title/Abstract] OR "Economic support" [Title/Abstract] OR "Financial support" [Title/Abstract]) AND ("2012"[Date - Publication] : "2021"[Date - Publication]) | 308 |
|  | (All TB treatment terms by title and abstract with OR as the Boolean operator) AND (All intervention terms by title and abstract with OR as the Boolean operator) AND ("2012"[Date - Publication] : "2021"[Date - Publication]) (i.e. #1 + additional terms) | 17,461 |
|  | (Tuberculosis[MeSH] + all TB treatment terms by title and abstract with OR as the Boolean operator) AND (All intervention terms by title and abstract with OR as the Boolean operator) AND ("2012"[Date - Publication] : "2021"[Date - Publication]) (i.e. #2+ MeSH terms) | 17,732 |
|  | ("2012"[Date - Publication] : "2021"[Date - Publication]) AND (Tuberculosis[MeSH] OR all TB treatment terms by title and abstract) AND (all intervention terms by title and abstract) AND (all outcome terms by title and abstract) (i.e. #3 + outcome terms). | 17,732 |

Web of Science search strategy
|  | Searches | Results |
| --- | --- | --- |
|  | TS=(TB terms with OR as the Boolean operator) AND TS=(all intervention terms with OR as the Boolean operator) AND TS=(all outcome terms with OR as the Boolean operator) | 28,985 |
|  | TI=(TB terms with OR as the Boolean operator) AND TS=(all intervention terms with OR as the Boolean operator) AND TS=(all outcome terms with OR as the Boolean operator) | 14,687 |
|  | TI=(TB terms with OR as the Boolean operator) AND TI=(all intervention terms with OR as the Boolean operator) AND TI=(all outcome terms with OR as the Boolean operator) | 1412 |
|  | TI=(TB terms with OR as the Boolean operator) AND TI=(all intervention terms with OR as the Boolean operator) AND TS=(all outcome terms with OR as the Boolean operator) | 3568 |

## REFERENCES

1. World Health Organization. Global Tuberculosis Report.; 2024. doi:978 92 4 156450 2

2. Boccia D, Rudgard W, Shrestha S, et al. Modelling the impact of social protection on tuberculosis: The S-PROTECT project. BMC Public Health. 2018;18(1):1–9. doi:10.1186/s12889-018-5539-x

3. Wingfield T, Tovar MA, Huff D, et al. The economic effects of supporting tuberculosis-affected households in Peru. Eur Respir J. 2016;48(5):1396–1410. doi:10.1183/13993003.00066-2016

4. Wingfield T, Boccia D, Tovar M, et al. Defining Catastrophic Costs and Comparing Their Importance for Adverse Tuberculosis Outcome with Multi-Drug Resistance: A Prospective Cohort Study, Peru. PLoS Med. 2014;11(7). doi:10.1371/journal.pmed.1001675

5. Bank TW. The World Bank In Social Protection. https://www.worldbank.org/en/topic/socialprotection/overview

6. Lönnroth K, Raviglione M. The WHO’s new end tb strategy in the post-2015 era of the sustainable development goals. Trans R Soc Trop Med Hyg. 2015;110(3):148–150. doi:10.1093/trstmh/trv108

7. Assembly UNG. Political Declaration of the High-Level Meeting on the Fight against Tuberculosis.; 2023.

8. Uplekar M, Weil D, Lonnroth K, et al. WHO’s new end TB strategy. Lancet. 2015;385(9979):1799–1801. doi:10.1016/S0140-6736(15)60570-0

9. Boccia D, Pedrazzoli D, Wingfield T, et al. Towards cash transfer interventions for tuberculosis prevention, care and control key operational challenges and research priorities. BMC Infect Dis. 2016;16. doi:10.1186/s12879-016-1529-8

10. Wingfield T, Tovar MA, Huff D, et al. A randomized controlled study of socioeconomic support to enhance tuberculosis prevention and treatment, Peru. Bull World HealthOrgan. 2017;95(4):270–280. doi:10.2471/BLT.16.170167

11. Richterman A, Steer-Massaro J, Jarolimova J, Nguyen LBL, Werdenberg J, Ivers LC. Cash interventions to improve clinical outcomes for pulmonary tuberculosis: Systematic review and meta-analysis. Bull World Health Organ. 2018;96(7):471–483. doi:10.2471/BLT.18.208959

12. Shamseer L, Moher D, Clarke M, Ghersi D, Liberati A, Petticrew M, Shekelle P SL. PRISMA-P (Preferred Reporting Items for Systematic review and Meta-Analysis Protocols) 2015 checklist: recommended items to address in a systematic review protocol. BMJ Br Med J. 2015;350:g7647.

13. Todd H, Hudson M, Grolmusova N, et al. Social Protection Interventions for TB-Affected HouseholdsC: A Scoping Review. Published online 2023:1–10. doi:10.4269/ajtmh.22-0470

14. Hudson M, Todd H, Nalugwa T, Boccia D, Wingfield T, Shete PB. The impact of social protection interventions on treatment and socioeconomic outcomes of people with tuberculosis and their households: Protocol for a systematic review and meta-analysis. Wellcome Open Res. 2023;8. doi:10.12688/wellcomeopenres.18807.1

15. Veritas Health Innovation. Covidence systematic review software.

16. Hamadeh, Nada, Van Rompaey, Catherine, Metreau E. World Bank Group country classifications by income level for FY24 (July 1, 2023-June 30, 2024). World Bank blogs. Published 2023. https://blogs.worldbank.org/en/opendata/new-world-bank-group-country-classifications-income-level-fy24

17. The World Bank. RESILIENCE, EQUITY, AND OPPORTUNITY The World Bank’s Social Protection and Labor Strategy 2012–2022.; 2022.

18. Ukwaja KN. Social protection interventions could improve tuberculosis treatment outcomes. LANCET Glob Heal. 2019;7(2):E167–E168. doi:10.1016/S2214-109X(18)30523-0

19. DerSimonian R, Laird N. Meta-analysis in clinical trials revisited. Contemp Clin Trials. 2015;45:139–145. doi:10.1016/j.cct.2015.09.002

20. StataCorp. Stata Statistical Software: Release 17. Published online 2021.

21. Sterne JAC, Savović J, Page MJ, et al. RoB 2: A revised tool for assessing risk of bias in randomised trials. BMJ. 2019;366(August). doi:10.1136/bmj.l4898

22. GA Wells, B Shea, D O’Connell, J Peterson, V Welch, M Losos, P Tugwell. The Newcastle-Ottawa Scale (NOS) for assessing the quality of nonrandomised studies in meta-analyses.

23. George LS, Rakesh PS, Sunilkumar M, Vijayakumar K, Kunoor A, Kumar V A. TB patient support systems in Kerala: A qualitative analysis. Indian J Tuberc. 2021;68(1):9–15. doi:10.1016/j.ijtb.2020.11.005

24. Kaliakbarova G, S. P, N. Z, G. R, B. T, S. van den H. Psychosocial support improves treatment adherence among MDR-TB patients: Experience from East Kazakhstan. Open Infect Dis J. 2013;7(SPEC ISS1):60–64. http://www.embase.com/search/results?subaction=viewrecord&from=export&id=L369141965 http://dx.doi.org/10.2174/1874279301307010060 http://findit.library.jhu.edu/resolve?sid=EMBASE&issn=18742793&id=doi:10.2174%2F1874279301307010060&atitle=Psychosocial

25. Orlandi GM, Pereira EG, Mineo Biagolini RE, de Siqueira Franca FO, Bertolozzi MR. Social incentives for adherence to tuberculosis treatment. Rev Bras Enferm. 2019;72(5):1182–1188. doi:10.1590/0034-7167-2017-0654

26. Ukwaja KN, Alobu I, Gidado M, Onazi O, Oshi DC. Economic support intervention improves tuberculosis treatment outcomes in rural Nigeria. Int J Tuberc Lung Dis. 2017;21(5):564–570. doi:10.5588/ijtld.16.0741

27. de Souza RA, Nery S, Rasella D, et al. Family health and conditional cash transfer in Brazil and its effect on tuberculosis mortality. Int J Tuberc LUNG Dis. 2018;22(11):1300+. doi:10.5588/ijtld.17.0907

28. Rudgard WE, das Chagas NS, Gayoso R, et al. Uptake of governmental social protection and financial hardship during drug-resistant tuberculosis treatment in Rio de Janeiro, Brazil. Eur Respir J. 2018;51(3). doi:10.1183/13993003.00274-2018

29. Wingfield T, Tovar MA, Huff D, et al. Beyond pills and tests: addressing the social determinants of tuberculosis. Clin Med. 2016;16(6):s79–s91. doi:10.7861/clinmedicine.16-6-s79

30. Florentino JL, Arao RML, Garfin AMC, et al. Expansion of social protection is necessary towards zero catastrophic costs due to TB: The first national TB patient cost survey in the Philippines. PLoS One. 2022;17(2 February):1–19. doi:10.1371/journal.pone.0264689

31. Pham TAM, Forse R, Codlin AJ, et al. Determinants of catastrophic costs among households affected by multi-drug resistant tuberculosis in Ho Chi Minh City, Viet Nam: a prospective cohort study. BMC Public Health. 2023;23(1):1–19. doi:10.1186/s12889-023-17078-5

32. Li R, Ruan Y, Sun Q, et al. Effect of a comprehensive programme to provide universal access to care for sputum-smear-positive multidrug-resistant tuberculosis in China: a before-and-after study. LANCET Glob Heal. 2015;3(4):E217–E228. doi:10.1016/S2214-109X(15)70021-5

33. Xiang L, Pan Y, Hou S, et al. The impact of the new cooperative medical scheme on financial burden of tuberculosis patients: evidence from six counties in China. Infect Dis Poverty. 2016;5(8).

34. Zhao Q, Wang L, Tao T, Xu B. Impacts of the “transport subsidy initiative on poor TB patients{’’} in Rural China: A Patient-Cohort Based Longitudinal Study in Rural China. PLoS One. 2013;8(11). doi:10.1371/journal.pone.0082503

35. Pedrazzoli D, Carter DJ, Borghi J, Laokri S, Boccia D, Houben RM. Does Ghana’s National Health Insurance Scheme provide financial protection to tuberculosis patients and their households? Soc Sci Med. 2021;277(March):113875. doi:10.1016/j.socscimed.2021.113875

36. Liu X, Lin KH, Li YH, et al. Impacts of Medical Security Level on Treatment Outcomes of Drug-Resistant Tuberculosis: Evidence from Wuhan City, China. Patient Prefer Adherence. 2022;16(December):3341–3355. doi:10.2147/PPA.S389231

37. Bhargava A, Bhargava M, Meher A, et al. Nutritional support for adult patients with microbiologically confirmed pulmonary tuberculosis: outcomes in a programmatic cohort nested within the RATIONS trial in Jharkhand, India. Lancet Glob Heal. 2023;11(9):e1402–e1411. doi:10.1016/S2214-109X(23)00324-8

38. Chenciner L, Annerstedt KS, Pescarini JM, Wingfield T. Social and health factors associated with unfavourable treatment outcome in adolescents and young adults with tuberculosis in Brazil: a national retrospective cohort study. Lancet Glob Heal. 2021;9(10):e1380–e1390. doi:10.1016/S2214-109X(21)00300-4

39. Dave JD, Rupani MP. Does Direct Benefit Transfer Improve Outcomes Among People With Tuberculosis? – A Mixed-Methods Study on the Need for a Review of the Cash Transfer Policy in India. Int J Heal Policy Manag. 2022;11(11):2552–2562. doi:10.34172/ijhpm.2022.5784

40. Randhawa KS, Khattak LU, Shaukat FA, Rafique S, Nasir M, Hayat S. Outcome Optimization For Patients With Drug-Resistant Tb Via The Implementation Of An All-Inclusive Care Program. J Pharm Negat Results. 2023;14(03):4001–4006. doi:10.47750/pnr.2023.14.03.504

41. Wrohan I, Nguyen TA, Nguyen VN, et al. Predictors of treatment outcomes among patients with multidrug-resistant tuberculosis in Vietnam: a retrospective cohort study. BMC Infect Dis. 2022;22(1):1–12. doi:10.1186/s12879-021-06992-x

42. Timire C, Sandy C, Ferrand RA, et al. Coverage and effectiveness of conditional cash transfer for people with drug resistant tuberculosis in Zimbabwe: A mixed methods study. PLOS Glob Public Heal. 2022;2(12):e0001027. doi:10.1371/journal.pgph.0001027

43. Higgins J, Savović J, Page MJ, Sterne JAC. RoB 2: A revised Cochrane risk-of-bias tool for randomized trials. Br Med J. 2019;(July):1–24. https://methods.cochrane.org/

44. Coding NS, For M, Studies C, et al. Appendix B Newcastle-Ottawa Scale Coding Manual. 2014;(Dc):19–20.

45. Ukwaja KN, Alobu I, Mustapha G, Onazi O, Oshi DC. ‘Sustaining the DOTS’: stakeholders’ experience of a social protection intervention for TB in Nigeria. Int Health. 2017;9(2):112–117. doi:10.1093/inthealth/ihx001

46. Xiang L, Pan Y, Hou S, et al. The impact of the new cooperative medical scheme on financial burden of tuberculosis patients: evidence from six counties in China. Infect Dis POVERTY. 2016;5. doi:10.1186/s40249-015-0094-5

47. de Andrade KV, Nery JS, de Souza RA, Pereira SM. Effects of social protection on tuberculosis treatment outcomes in low or middle-income and in high-burden countries: systematic review and meta-analysis. Cad Saude Publica. 2018;34(1). doi:10.1590/0102-311X00153116

48. World Health Organization (WHO). Guidance on social protection for people affected by tuberculosis. Language (Baltim). Published online 2024:1–13.

49. Organization IL. Topic portal: Social protection.

50. Hargreaves JR, Boccia D, Evans CA, Adato M, Petticrew M. The Social Determinants of TuberculosisC: From Evidence to Action. 2011;101(4):654–662. doi:10.2105/AJPH.2010.199505

