## Supplementary table 1-6 for "The impact of social protection interventions on treatment and socioeconomic outcomes of tuberculosis-affected people and households in low income, high burden settings: A systematic review and meta-analysis"

### **Supplementary Table 1.** Study characteristics of quantitative studies (n=46)

| **Authors** | **Study Title** | **Year of Publication** | **Country** | **Setting** |
| --- | --- | --- | --- | --- |
| Bhargava  et al. | Nutritional support for adult patients with microbiologically confirmed pulmonary tuberculosis: outcomes in a programmatic cohort nested within the RATIONS trial in Jharkland, India | 2023 | India | Rural |
| Bhatt R  et al. | Impact of integrated psycho-socio-economic support on treatment outcome in drug resistant tuberculosis - A retrospective cohort study. | 2019 | India | Urban |
| Carter  et al. | The impact of a cash transfer programme on tuberculosis treatment success rate: a quasi-experimental study in Brazil. | 2019 | Brazil | Mixed |
| Chenicer  et al. | Social and health factors associated with unfavourable treatment outcome in adolescents and young adults with tuberculosis in Brazil: a national retrospective cohort study | 2021 | Brazil | Mixed |
| Ciobanu  et al. | Do incentives improve tuberculosis treatment outcomes in the Republic of Moldova? | 2014 | Moldova | Mixed |
| Dave et al. | Does Direct Benefit Transfer Improve Outcomes Among People With Tuberculosis? – A Mixed-Methods Study on the Need for a Review of the Cash Transfer Policy in India | 2022 | India | Mixed |
| de Souza  et al. | Family health and conditional cash transfer in Brazil and its effect on tuberculosis mortality. | 2018 | Brazil | Mixed |
| Durovni  et al. | The impact of the Brazilian family health strategy and the conditional cash transfer on tuberculosis treatment outcomes in Rio De Janeiro: an individual-level analysis of secondary data | 2018 | Brazil | Urban |
| Florentino et al. | Expansion of social protection is necessary towards zero catastrophic costs due to TB: The first national TB patient cost survey in the Philippines | 2022 | Philippines | Mixed |
| Jeyashree et al. | Ni-kshay Poshan Yojana: receipt and utilization among persons with TB notified under the National TB Elimination Program in India, 2022 | 2024 | India | Mixed |
| Jiang et al. | Factors associated with loss to follow-up before and after treatment initiation among patients with tuberculosis: A 5-year observation in China | 2023 | China | Not specified |
| Klein et al. | Evaluation of a social protection policy on tuberculosis treatment outcomes: A prospective cohort study | 2019 | Argentina | Urban |
| Li et al. | Effect of a comprehensive programme to provide universal access to care for sputum-smear-positive multidrug-resistant tuberculosis in China: a before-and-after study. | 2015 | China | Urban |
| Liu et al. | Impacts of Medical Security Level on Treatment Outcomes of Drug-Resistant Tuberculosis: Evidence from Wuhan City, China | 2023 | China | Urban |
| Lutge et al. | Economic support to improve tuberculosis treatment outcomes in South Africa: a pragmatic cluster-randomized controlled trial. | 2013 | South Africa | Mixed |
| Mahapatra et al. | Effectiveness of food supplement on treatment outcomes and quality of life in pulmonary tuberculosis: Phased implementation approach | 2024 | India | Mixed |
| Mansour  et al. | Impact of a nutritional support programme on loss to follow-up after tuberculosis diagnosis in Kenya | 2018 | Kenya | Mixed |
| Modi  et al. | Financial Incentive - does this have an impact on outcome of Tuberculosis? | 2020 | India | Urban |
| Ngamvithay apong-Yanai et al. | Engaging women volunteers of high socioeconomic status in supporting socioeconomically disadvantaged tuberculosis patients in Chiang Rai, Thailand. | 2013 | Thailand | Urban |
| Oliosi et al. | Effect of the Bolsa Familia Programme on the outcome of tuberculosis treatment: a prospective cohort study. | 2019 | Brazil | Urban |
| Pavinati et al. | Vulnerability to loss of follow-up and death due to tuberculosis among homeless individuals in Brazil: a retrospective cohort study | 2024 | Brazil | Not specified |
| Pedrazzoli et al. | Does Ghana’s National Health Insurance Scheme provide financial protection to tuberculosis patients and their households? | 2021 | Ghana | Mixed |
| Pham et al. | Determinants of catastrophic costs among households affected by multi-drug resistant tuberculosis in Ho Chi Minh City, Viet Nam: a prospective cohort study | 2023 | Vietnam | Urban |
| Potty et al. | Tuberculosis treatment outcomes and patient support groups, southern India | 2023 | India | Urban |
| Priedeman et al. | Evaluating the impact of social support services on tuberculosis treatment default in Ukraine | 2018 | Ukraine | Mixed |
| Randhawa et al. | Outcome Optimization for Patients with Drug-Resistant TB Via The Implementation of An All-Inclusive Care Program | 2023 | Pakistan | Not specified |
| Reis-Santos et al. | A Matter of Inclusion: A Cluster-Randomized Trial to Access the Effect of Food Vouchers Versus Traditional Treatment on Tuberculosis Outcomes in Brazil | 2022 | Brazil | Urban |
| Reis-Santos et al. | Tuberculosis in Brazil and cash transfer programs: A longitudinal database study of the effect of cash transfer on cure rates. | 2019 | Brazil | Mixed |
| Rogers  et al. | Impact of community-based adherence support on treatment outcomes for tuberculosis, leprosy and HIV/AIDS-infected individuals in post-Ebola Liberia. | 2018 | Liberia | Not specified |
| Rohit et al. | Does provision of cash incentive to HIV-infected tuberculosis patients improve the treatment success in programme settings? A cohort study from South India. | 2020 | India | Mixed |
| Rudgard  et al. | Uptake of governmental social protection and financial hardship during drug-resistant tuberculosis treatment in Rio de Janeiro, Brazil. | 2017 | Brazil | Urban |
| Samuel  et al. | Relationship between nutritional support and tuberculosis treatment outcomes in West Bengal India | 2015 | India | Rural |
| Singh et al. | Improving tuberculosis treatment success rate through nutrition supplements and counseling: Findings from a pilot intervention in India | 2021 | India | Not specified |
| Soares et al. | Tuberculosis control in a socially vulnerable area: a community intervention beyond DOT in a Brazilian favela. | 2013 | Brazil | Urban |
| Sripad et al. | Effects of Ecuador's national monetary incentive program on adherence to treatment for drug-resistant tuberculosis. | 2014 | Ecuador | Mixed |
| Timire et al. | Coverage and effectiveness of conditional cash transfer for people with drug resistant tuberculosis in Zimbabwe: A mixed methods study | 2022 | Zimbabwe | Mixed |
| Torrens  et al. | Effectiveness of a conditional cash transfer programme on TB cure rate: A retrospective cohort study in Brazil | 2016 | Brazil | Mixed |
| Ukwaja  et al. | Economic Support Intervention improves tuberculosis treatment outcomes in Rural Nigeria | 2017 | Nigeria | Rural |
| Wei et al. | Providing financial incentives to rural-to-urban tuberculosis migrants in Shanghai: an intervention study. | 2012 | China | Urban |
| Wingfield et al. | Beyond pills and tests: addressing the social determinants of tuberculosis | 2016 | Peru | Urban |
| Wingfield et al. | The economic effects of supporting tuberculosis-affected households in Peru. | 2016 | Peru | Urban |
| Wingfield et al. | A randomized controlled study of socioeconomic support to enhance tuberculosis prevention and treatment, Peru. | 2017 | Peru | Urban |
| Wrohan  et al. | Predictors of treatment outcomes among patients with multidrug-resistant tuberculosis in Vietnam: a retrospective cohort study | 2022 | Vietnam | Not specified |
| Xiang et al. | The impact of the new cooperative medical scheme on financial burden of tuberculosis patients: evidence from six counties in China. | 2016 | China | Urban |
| Yin et al. | The relationship between social support, treatment interruption and treatment outcome in patients with multidrug resistant tuberculosis in China: A mixed methods study. | 2018 | China | Urban |
| Zhao et al. | Impacts of the "transport subsidy initiative on poor TB patients" in Rural China: a patient-cohort based longitudinal study in rural china | 2013 | China | Rural |

### **Supplementary Table 2.** Study characteristics of qualitative studies (n=4)

| **Authors** | **Study Title** | **Year of Publication** | **Country** | **Setting** |
| --- | --- | --- | --- | --- |
| George et al. | TB patient support systems in Kerala: A qualitative analysis. | 2021 | India | Urban |
| Kaliakbarova et al. | Psychosocial support improves treatment adherence among MDR-TB patients: Experience from East Kazakhstan | 2013 | Kazakhst-an | Mixed, primarily urban |
| Orlandi et al. | Social incentives for adherence to tuberculosis treatment | 2019 | Brazil | Urban |
| Ukwaja et al. | Sustaining the DOTS': stakeholders' experience of a social protection intervention for TB in Nigeria | 2021 | Nigeria | Rural |

### **Supplementary Table 3.** Social protection interventions described in quantitative studies (n=46)

| **Study** | **Primary funding source** | **Social protection intervention type** | **Eligibility for social**  **protection intervention** | **TB sensitive vs. TB specific** |
| --- | --- | --- | --- | --- |
| Bhargava et al. | Study intervention/source | Nutritional support | Not described | Not described |
| Bhatt R et al. | Government/national TB program | Mixed with psychosocial support | Not described | Not described |
| Carter et al. | Government/national TB program | Cash | Not described | Not described |
| Chenicer  et al. | Government/national TB program | Cash | Measures of poverty | TB sensitive |
| Ciobanu et al. | Mixed funding sources | Mixed without psychosocial support | TB status | TB sensitive |
| Dave et al. | Government/ national TB program | Cash | TB status | TB specific |
| de Souza et al. | Government/ national TB program | Cash | Measures of poverty | TB sensitive |
| Durovni et al. | Government/ national TB program | Cash | Measures of poverty | TB sensitive |
| Florentino et al. | Government/ national TB program | Mixed without psychosocial support | TB status | TB specific |
| Jeyashree et al. | Government/national TB program | Cash | TB status | TB specific |
| Jiang et al. | Government/national TB program | Social health insurance | Not described | TB sensitive |
| Klein et al. | Government/national TB program | Cash | TB status | TB sensitive |
| Li et al. | Government/national TB program | Cash | TB status | TB sensitive |
| Liu et al. | Government | Social health insurance | Not described | TB sensitive |
| Lutge et al | Study intervention/source | Cash | Study intervention | TB specific |
| Mahapatra et al. | Study intervention/source | Nutritional support | Study intervention | TB specific |
| Mansour et al. | Government/ national TB program | Nutritional support | TB status | TB sensitive |
| Modi et al. | Government/ national TB program | Cash | TB status | TB specific |
| Ngamvithay-apong-Yanai et al. | Study intervention/source | Mixed with psychosocial support | Study intervention | TB specific |
| Oliosi et al. | Government/ national TB program | Cash | Measures of poverty | TB sensitive |
| Pavinati et al. | Government/national TB program | Cash | Not described | Not described |
| Pedrazzoli et al. | Government/ national TB program | Social health insurance | Not described | TB sensitive |
| Pham et al. | Government/ national TB program | Mixed without psychosocial support | TB status | TB speciific |
| Potty et al. | Study intervention/source | Psychosocial support | TB status | TB specific |
| Priedeman et al. | NGO/multilateral agency | Mixed without psychosocial support | TB status | TB sensitive |
| Randhawa et al. | Study intervention/source | Mixed with psychosocial support | TB status | TB specific |
| Reis-Santos et al (2022) | Study intervention/source | Nutritional support | Study intervention | TB specific |
| Reis-Santos et al. (2019) | Government/ national TB program | Cash | Measures of poverty | TB sensitive |
| Rogers et al. | NGO/multilateral agency | Mixed with psychosocial support | Study intervention | TB specific |
| Rohit et al. | Government/ national TB program | Cash | TB status | TB sensitive |
| Rudgard et al. | Government/ national TB program | Cash | Measures of poverty | TB sensitive |
| Samuel et al. | NGO/multilateral agency | Nutritional support | TB status | TB sensitive |
| Singh et al. | Study intervention/source | Nutritional support | TB status | TB specific |
| Soares et al. | Government/national TB program | Primarily psychosocial support | TB status | TB sensitive |
| Sripad et al. | Government/national TB program | Cash | TB status | TB sensitive |
| Timire et al. | Government/national TB program | Cash | TB status | TB specific |
| Torrens et al. | Government/national TB program | Cash | Measures of poverty | TB sensitive |
| Ukwaja et al. | Study intervention/source | Cash | TB status | TB sensitive |
| Wei et al. | Study intervention/source | Cash | Study intervention | TB specific |
| Wingfield et al. (2016a) | Study intervention/source | Mixed with psychosocial support | Study intervention | TB specific |
| Wingfield et al. (2016b) | Study intervention/source | Mixed with psychosocial support | Study intervention | TB specific |
| Wingfield et al. (2017) | Study intervention/source | Mixed with psychosocial support | Study intervention | TB specific |
| Wrohan et al. | Government/national TB program | Social health insurance | Not described | TB sensitive |
| Xiang et al. | Government/national TB program | Cash | Other | TB specific |
| Yin et al. | Not specified | Mixed with psychosocial support | Study intervention | TB specific |
| Zhao et al. | NGO/multilateral agency | Cash | TB status | TB specific |

### **Supplementary Table 4.** Outcomes and implementation challenges described in quantitative studies (n=46). “Mix of other TB treatment outcomes” include mortality, TB treatment default (treatment interruption of at least two months), loss to follow up, or treatment failure.

| **Study** | **Primary outcome** | **Secondary outcome** | **Primary implementation challenge** | **Secondary implementation challenge** |
| --- | --- | --- | --- | --- |
| Bhargava et al. | TB treatment success | Mix of other TB treatment outcomes | Not described | Not described |
| Bhatt R et al. | TB treatment success | Mix of other TB treatment outcomes | Not described | Not described |
| Carter et al. | TB treatment success | N/A | Not described | Not described |
| Chenicer et | Unfavorable treatment outcomes | Mix of other TB treatment outcomes | Not described | Not described |
| Ciobanu et al. | TB treatment success | Mix of other TB treatment outcomes | Not described | Not described |
| Dave et al. | Unfavorable treatment outcomes | TB treatment success | Delayed payments or non-receipt of payment | Insufficient funds |
| de Souza et al. | Death | N/A | Not described | Not described |
| Durovni et al. | TB treatment success | N/A | Not described | Not described |
| Florentino et al. | Catastrophic costs and dissaving | Financial coping mechanisms | Not described | Not described |
| Jeyashree et al. | TB treatment success | Mix of other TB treatment outcomes | Delayed payments or non-receipt of payment | Not described |
| Jiang et al. | Loss to follow up | N/A | Not described | Not described |
| Klein et al. | TB treatment success | Mix of other TB treatment outcomes | Administrative issues (inadequate banking systems/issues with paperwork or forms required) | Lack of knowledge/awareness about the program or how to link patients to the program (patient or provider/provisioner) |
| Li et al. | Other positive TB treatment outcomes | Socioeconomic outcome | Not described | Not described |
| Liu et al. | TB treatment success | Out of pocket expenses | Not described | Not described |
| Lutge et al. | TB treatment success | N/A | Administrative issues (inadequate banking systems/issues with paperwork or forms required, delayed disbursement or non-receipt of funds/benefit) | Not described |
| Mahapatra et al. | TB treatment success | Mix of other TB treatment outcomes | Not described | Not described |
| Mansour et al. | Loss to follow up | N/A | Not described | Not described |
| Modi et al. | TB treatment success | Mix of other TB treatment outcomes | Administrative issues (inadequate banking systems/issues with paperwork or forms required, delayed disbursement or non-receipt of funds/benefit) | Lack of knowledge/awareness about the program or how to link patients to the program (patient or provider/provisioner) |
| Ngamvithay-apong-Yanai et al. | TB treatment success | Treatment failure | Not described | Not described |
| Oliosi et al. | TB treatment success | Mix of other TB treatment outcomes | Not described | Not described |
| Pavinati et al. | Unfavorable treatment outcomes | Cure | Not described | Not described |
| Pedrazzoli et al. | Catastrophic costs | Direct and indirect costs | Not described | Not described |
| Pham et al. | Catastrophic costs | Household income, income loss | Not described | Not described |
| Potty et al. | TB treatment success | N/A | Not described | Not described |
| Priedeman et al. | TB treatment success | Mix of other TB treatment outcomes | Not described | Not described |
| Randhawa et al. | TB treatment success | Mix of other TB treatment outcomes | Not described | Not described |
| Reis-Santos et al. (2022) | TB treatment success | Nutritional support | Not described | Not described |
| Reis-Santos et al. (2019) | TB treatment success | N/A | Not described | Not described |
| Rogers et al. | TB treatment success | Mix of other TB treatment outcomes | Not described | Not described |
| Rohit et al. | TB treatment success | Mix of other TB treatment outcomes | Administrative issues (inadequate banking systems/issues with paperwork or forms required, delayed disbursement or non-receipt of funds/benefit) | Lack of knowledge/awareness about the program or how to link patients to the program (patient or provider/provisioner) |
| Rudgard et al | Socioeconomic outcome | N/A | Not described | Not described |
| Samuel et al. | Unsuccessful treatment outcome | Mix of other TB treatment outcomes | Not described | Not described |
| Singh et al. | Treatment success | Weight gain | Not described | Not described |
| Soares et al. | TB treatment success | Mix of other TB treatment outcomes | Not described | Not described |
| Sripad et al. | Treatment default | N/A | Administrative issues (inadequate banking systems/issues with paperwork or forms required) | Not described |
| Timire et al. | TB treatment success | Mix of other TB treatment outcomes | Administrative issues (inadequate banking systems/issues with paperwork or forms required, delayed disbursement or non-receipt of funds/benefit), insufficient funds | Lack of knowledge/awareness about the program or how to link patients to the program (patient or provider/provisioner) |
| Torrens et al. | TB treatment success | N/A | Not described | Not described |
| Ukwaja et al | TB treatment success | Mix of other TB treatment outcomes | Not described | Not described |
| Wei et al. | TB treatment success | Mix of other TB treatment outcomes | Administrative issues (inadequate banking systems/issues with paperwork or forms required) | Not described |
| Wingfield et al. (2016a) | Socioeconomic outcome | N/A | Not described | Not described |
| Wingfield et al. (2016b) | Socioeconomic outcome | N/A | Not described | Not described |
| Wingfield et al. (2017) | TB treatment success | Mix of other TB treatment outcomes | Not described | Not described |
| Wrohan et al | Loss to follow up | TB treatment success | Not described | Not described |
| Xiang et al. | Socioeconomic outcome | N/A | Not described | Not described |
| Yin et al. | TB treatment success | Mix of other TB treatment outcomes | Not described | Not described |
| Zhao et al. | Socioeconomic outcome | N/A | Other-insufficient funds | Not described |

### **Supplementary Table 5.** Risk of bias assessments of non-randomized studies using the Newcastle Ottawa Scale (NOS).

| **Study** | **Selection (maximum score=4)** | **Comparability**  **(maximum score=2)** | **Outcome (maximum score=3)** |
| --- | --- | --- | --- |
| Bhargava, 2023 | 4 | 2 | 3 |
| Bhatt et al., 2019 | 3 | 0 | 3 |
| Carter et al., 2019 | 4 | 2 | 3 |
| Chenicer et al. | 4 | 2 | 3 |
| Ciobanu et al., 2014 | 4 | 2 | 3 |
| Dave et al., 2022 | 4 | 2 | 3 |
| de Souza et al., 2018 | 4 | 2 | 3 |
| Durovni et al., 2018 | 4 | 2 | 3 |
| Florentino et al. 2022 | 4 | 2 | 3 |
| Jeyashree et al., 2024 | 4 | 2 | 3 |
| Jiang et al. 2023 | 4 | 2 | 3 |
| Klein et al., 2019 | 4 | 2 | 3 |
| Li et al., 2015 | 4 | 2 | 3 |
| Liu et al., 2023 | 4 | 2 | 3 |
| Mahapatra et al. | 4 | 2 | 3 |
| Mansour et al., 2018 | 4 | 2 | 3 |
| Modi et al., 2020 | 3 | 0 | 2 |
| Ngamvithayapong-Yanai et al., 2013 | 3 | 1 | 3 |
| Oliosi et al., 2019 | 4 | 2 | 3 |
| Pavinati et al., 2024 | 4 | 2 | 2 |
| Pedrazzoli et al., 2021 | 4 | 2 | 3 |
| Potty et al., 2023 | 4 | 2 | 3 |
| Priedeman et al., 2018 | 4 | 2 | 3 |
| Randhawa et al., 2023 | 1 | 0 | 1 |
| Reis-Santos et al., 2019 | 4 | 2 | 3 |
| Rogers et al., 2018 | 4 | 2 | 3 |
| Rohit et al., 2020 | 4 | 2 | 3 |
| Rudgard et al., 2018 | 4 | 2 | 3 |
| Samuel et al., 2016 | 4 | 1 | 3 |
| Singh et al., 2021 | 4 | 1 | 3 |
| Soares et al., 2013 | 4 | 2 | 3 |
| Sripad et al., 2014 | 4 | 2 | 3 |
| Timire et al., 2022 | 4 | 2 | 3 |
| Torrens et al., 2016 | 4 | 2 | 3 |
| Ukwaja et al., 2017 | 4 | 2 | 3 |
| Wei et al., 2012 | 4 | 2 | 3 |
| Wrohan et al., 2022 | 4 | 2 | 3 |
| Xiang et al., 2016 | 4 | 2 | 3 |
| Yin et al., 2018 | 1 | 0 | 0 |
| Zhao et al., 2013 | 4 | 2 | 3 |

### **Supplementary Table 6.** Risk of bias assessments of randomized controlled trials (RCTs) using the Cochrane Risk of Bias (RoB) assessment tool.

| **Study** | **Overall risk assessment** |
| --- | --- |
| Lutge et al., 2013 | Low risk |
| Pham et al., 2023 | Low risk |
| Reis-Santos et al. 2022 | Low risk |
| Wingfield et al., 2016 | Low risk |
| Wingfield et al., 2016 | Low risk |
| Wingfield et al., 2017 | Low risk |
