## Supplementary table 7 for "The impact of social protection interventions on treatment and socioeconomic outcomes of tuberculosis-affected people and households in low income, high burden settings: A systematic review and meta-analysis"

**Supplementary Table 7.** A listing of excluded studies (n=36)

| **First author** | **Year of publication** | **Title** | **Reason excluded** |
| --- | --- | --- | --- |
| Aderno et al. | 2012 | Support and social care of tuberculosis in pediatrics | Ineligible study design |
| [Aragão](https://pubmed.ncbi.nlm.nih.gov/?term=Arag%C3%A3o+FBA&cauthor_id=38896660) et al. | 2024 | Social protection in areas vulnerable to tuberculosis: a mixed methods study in São Luís, Maranhão | Ineligible outcomes |
| Amon et al. | 2022 | The impact of scaling up human rights interventions on reducing inequality and increasing access to care and treatment for HIV and TB: Mid-term results from the Breaking Down Barriers initiative | Ineligible study design |
| Balakrishnan et al. | 2015 | Social inclusion: An effort to end loss-to-treatment follow-up in tuberculosis | Ineligible intervention |
| Begum et al. | 2020 | Utilisation of nutritional support scheme among the patients of tuberculosis: A myth or a truth | Ineligible outcome |
| Cassidy et al. | 2018 | Food Insecurity and Tuberculosis: Policy Urgently Needs To Play Catch-Up | Ineligible outcome |
| Cavalcanti et al. | 2023 | Evaluation and Forecasting Analysis of the Association of Conditional Cash Transfer With Child Mortality in Latin America, 2000-2030 | Ineligible time frame |
| Claros et al. | 2014 | Adherence to HIV and TB care and treatment, the role of food security and nutrition | Ineligible outcome |
| Cocozza et al. | 2020 | An assessment of current tuberculosis patient care and support policies in high-burden countries | Ineligible outcome |
| Contreras et al. | 2017 | Addressing tuberculosis patients' medical and socio-economic needs: a comprehensive programmatic approach | Ineligible outcome |
| dePaulaMartins et al. | 2019 | Effect of the Bolsa Familiia Programme on tuberculosis treatment outcomes | Ineligible study design |
| Deshmukh et al. | 2018 | Social support a key factor for adherence to multidrug-resistant tuberculosis treatment | Ineligible outcome |
| Diaw et al. | 2018 | Implementing TB control in a rural, resource-limited setting: the stop-TB Italia project in Senegal | Ineligible intervention |
| Elias et al. | 2015 | Material benefits as an incentive to reduce abandonment of treatment of tuberculosis in people living in street: review of economic analysis | Ineligible outcome |
| Hu et al. | 2021 | Evaluation of the effect of the capitation compensation mechanism among pulmonary tuberculosis patients with a full period of treatment | Ineligible outcome |
| Kim et al. | 2018 | Tuberculosis Relief Belt Supporting Project (Tuberculosis Patient Management Project for Poverty Group) | Ineligible intervention |
| Klein et al. | 2019 | Correction: Evaluation of a social protection policy on tuberculosis treatment outcomes: A prospective cohort study | Ineligible study design |
| Kliner et al. | 2015 | Effects of financial incentives for treatment supporters on tuberculosis treatment outcomes in Swaziland: a pragmatic interventional study | Ineligible population |
| Kumar et al. | 2020 | Nikshay Poshan Yojana (NPY) for tuberculosis patients: Early implementation challenges in Delhi, India | Ineligible outcome |
| Lee et al. | 2019 | Incentives that influence low income Filipinos with Tuberculosis symptoms to change health-seeking behaviour: a randomized controlled trial | Ineligible outcome |
| Li et al. | 2015 | Effect of a comprehensive programme to provide universal access to care for sputum-smear-positive multidrug-resistant tuberculosis in China: a before-and-after study | Ineligible study design |
| Lönnroth et al. | 2014 | Beyond UHC: monitoring health and social protection coverage in the context of tuberculosis care and prevention. | Ineligible study design |
| Malacarne et al. | 2018 | Factors associated with TB in an indigenous population in Brazil: the effect of a cash transfer program | Ineligible outcome |
| Nery et al. | 2017 | Effect of Brazil's conditional cash transfer programme on tuberculosis incidence | Ineligible outcome |
| Nidoi et al. | 2021 | Impact of socio-economic factors on Tuberculosis treatment outcomes in north-eastern Uganda: a mixed methods study | Ineligible intervention |
| Parthasarathy et al. | 2019 | Cash transfer interventions in tuberculosis treatment outcomes | Ineligible study design |
| Patel et al. | 2019 | Cash transfer scheme for people with tuberculosis treated by the National TB Programme in Western India: a mixed methods study | Ineligible outcomes |
| Shah et al. | 2023 | Notes for Challenges and Strategic Solutions to Guarantee Last Mile Reach for an Indian TB Patient’s Nikshay Poshan Yojana; A Conditional Cash Transfer Scheme comment on “Does Direct Benefit Transfer Improve Outcomes Among People With Tuberculosiis? A Mixed-Method Study on the Need for a Review of the Cash Transfer Policy in India | Ineligible outcomes |
| Shah et al. | 2023 | Challenges and Strategic Solutions to Guarantee Last Mile Reach for an Indian TB Patient’s Nikshay Poshan Yojana; A Conditional Cash Transfer Scheme; Comment on “Does Direct Benefit Transfer Improve Outcomes Among People With Tuberculosis? – A Mixed-Methods Study on the Need for a Review of the Cash Transfer Policy in India” | Ineligible study design |
| Siroka et al. | 2016 | Association between spending on social protection and tuberculosis burden: a global analysis | Ineligible outcome |
| Suwannakeeree et al. | 2015 | A Medication Adherence Enhancement Program for Persons with Pulmonary Tuberculosis: A Randomized Controlled Trial Study. Pacific Rim International Journal of Nursing Research | Ineligible intervention |
| Testov et al. | 2014 | Impact of social support programme on MDR-TB patients' treatment outcomes | Ineligible study design |
| Traore et al. | 2022 | The high costs facing TB-affected households in Mali | Ineligible intervention |
| Vanleeuw et al. | 2022 | I’m suffering for food”: Food insecurity and access to social protection for TB patients and their households in Cape Town, South Africa | Ineligible outcome |
| Wei et al. | 2015 | Effective reimbursement rates of the rural health insurance among uncomplicated tuberculosis patients in China | Ineligible intervention |
| Wingfield et al. | 2015 | Designing and implementing a socioeconomic intervention to enhance TB control: operational evidence from the CRESIPT project in Peru. BMC Public Healt | Ineligible study design |
